# High-dimensional causal mediation analysis by partial sum statistic and sample splitting strategy in imaging genetics application

**DOI:** 10.1101/2024.06.23.24309362

**Authors:** Chang Hung-Ching, Fang Yusi, Michael T. Gorczyca, Batmanghelich Kayhan, George C. Tseng

## Abstract

Causal mediation analysis provides a systematic approach to explore the causal role of one or more mediators in the association between exposure and outcome. In omics or imaging data analysis, mediators are often high-dimensional, which brings new statistical challenges. Existing methods either violate causal assumptions or fail in interpretable variable selection. Additionally, mediators are often highly correlated, presenting difficulties in selecting and prioritizing top mediators. To address these issues, we develop a framework using Partial Sum Statistic and Sample Splitting Strategy, namely PS5, for high-dimensional causal mediation analysis. The method provides a powerful global mediation test satisfying causal assumptions, followed by an algorithm to select and prioritize active mediators with quantification of individual mediation contributions. We demonstrate its accurate type I error control, superior statistical power, reduced bias in mediation effect estimation, and accurate mediator selection using extensive simulations of varying levels of effect size, signal sparsity, and mediator correlations. Finally, we apply PS5 to an imaging genetics dataset of chronic obstructive pulmonary disease (COPD) patients (*N*=8,897) in the COPDGene study to examine the causal mediation role of lung images (*p*=5,810) in the associations between polygenic risk score and lung function and between smoking exposure and lung function, respectively. Both causal mediation analyses successfully estimate the global indirect effect and detect mediating image regions. Collectively, we find a region in the lower lobe of the right lung with a strong and concordant mediation effect for both genetic and environmental exposures. This suggests that targeted treatment toward this region might mitigate the severity of COPD due to genetic and smoking effects.

## 1. Introduction

Mediation analysis investigates the causal role of one or multiple mediators through which an exposure influences the outcome. In studies with omics or imaging data, mediators of interest are often high-dimensional, and an increasing number of methods have been developed for this purpose in the past few years (Zeng, Shao and Zhou, 2021). Our motivating example comes from an imaging genetics dataset from the Chronic Obstructive Pulmonary Disease Genetic Epidemiology (COPDGene) study (*N*=8,897) (Regan et al., 2011). We are interested in investigating computed tomography (CT) imaging as the potential causal mediator in the impact of polygenic risk score (PRS) on the lung functional outcome measured by forced expiratory volume (FEV1) (Figure 1A). More disease background and study information will be discussed in detail in Section 5. Unlike most neuroimaging genetic studies that summarize images into selected low-dimensional morphological or biological features in pre-specified regions of interest (ROI), we apply a deep learning algorithm (Li, Ke and Kayhan, 2021; Yu et al., 2024), an in-house self-supervised representation learning method, to extract *p* = 5, 810 features representing 581 local images (patches). The goal is to detect 3D physical locations in the lung that causally mediate the genetic effect (i.e., PRS) on lung functional outcome (i.e., FEV1). Specifically, three primary research aims are pursued: (A1) to conduct a powerful statistical test for detecting the global mediation (indirect) effect through lung imaging while satisfying causal assumptions; (A2) to quantify the amount (percentage) of global mediation effect; (A3) to prioritize 3D locations in lung as top active mediators and to quantify their mediation contributions.

**Fig 1:**
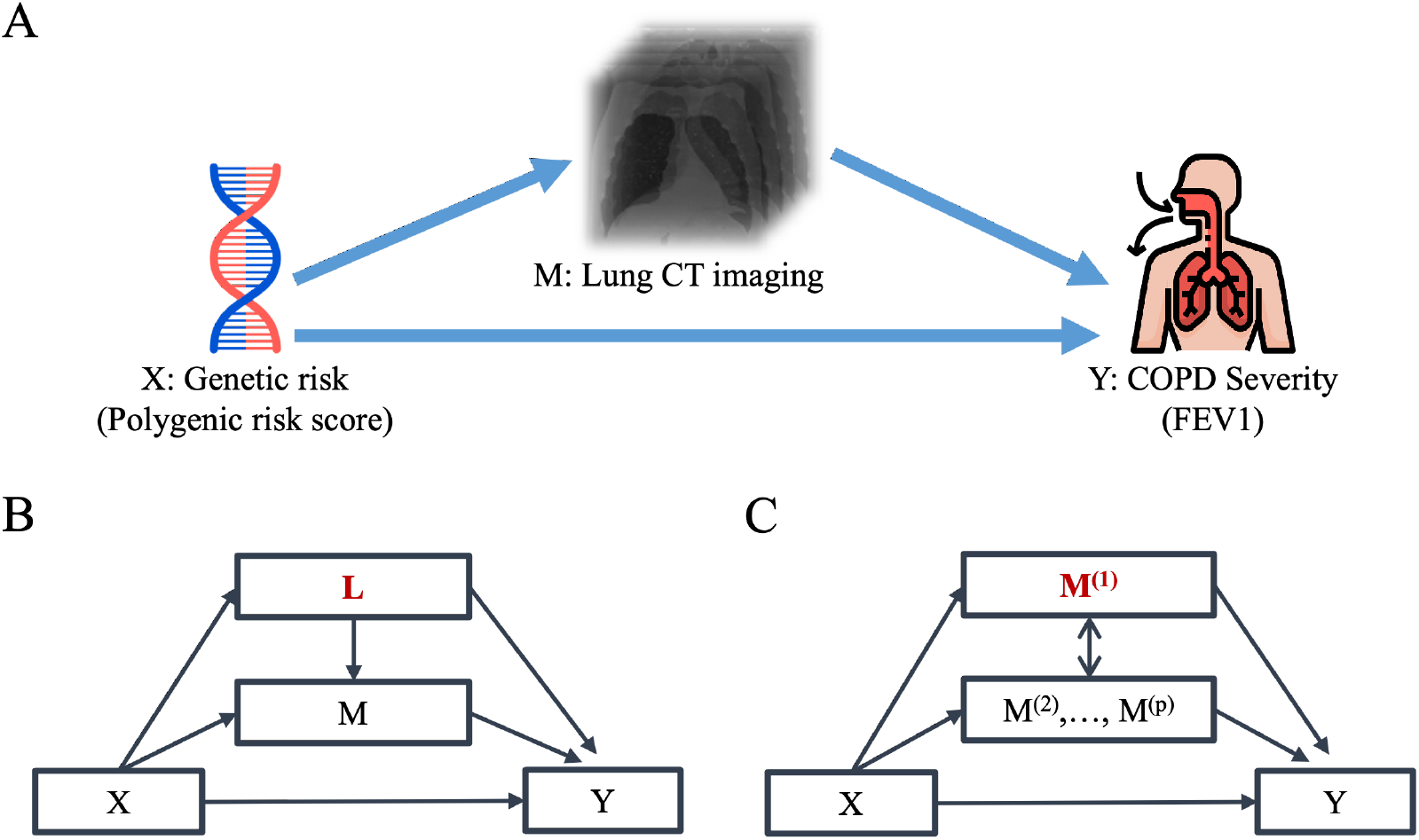
(A) Proposed causal pathway for COPD. (B) Example of *X*-induced confounder for the mediators-outcome relationship. (C) Example of potential *X*-induced confounder among high-dimensional mediators.

**Fig 2:**
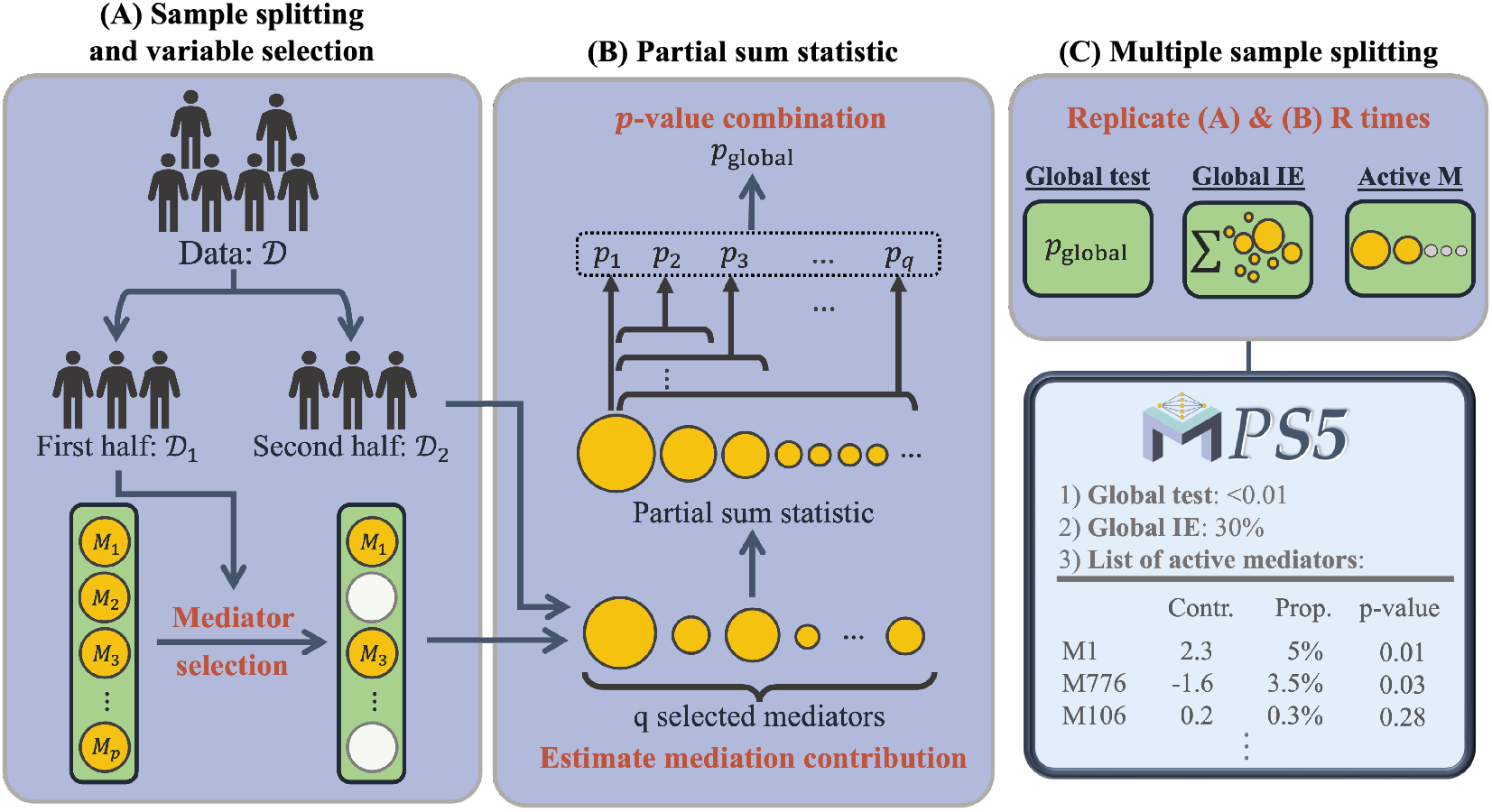
Graphical abstract of PS5, a three-step analysis framework including (A) Sample splitting and variable selection, (B) Partial sum statistic for testing global indirect effect, and (C) Multiple sample splitting and prioritization of selected mediators.

Although many methods have been developed, to our knowledge, no existing method provides a statistically rigorous framework for all aims (A1)-(A3). Existing methods often perform dimension reduction or variable selection prior to mediation analysis to detect mediation effects. These methods can be categorized into two groups based on their dimensionality reduction approaches: penalized regression (Zhang et al., 2016; Zhou, Wang and Zhao, 2020) and orthogonal transformation (Huang and Pan, 2016; Zhao, Lindquist and Caffo, 2020). The former category uses penalized regression or sparse priors to reduce dimensionality of mediators, allowing better interpretability. Despite their success, these methods do not explicitly verify causal assumptions in the dimension reduction and are limited in the analytical goals. For example, HILMA by Zhou, Wang and Zhao (2020) cannot prioritize mediator (aim A3), while HIMA by Zhang et al. (2016) does not conduct a statistical test for global mediation effect (aim A1). On the other hand, the latter category orthogonally transforms mediators to be uncorrelated and fits a series of single mediator models to ensure causal assumptions. Yet, these methods sacrifice interpretability, which is crucial in biological and clinical impact (Caruana et al., 2015), and have difficulties in prioritizing mediators (aim A3) since each transformed mediator is a linear combination of the original mediators. Moreover, as active mediators are relatively sparse, orthogonal transformation methods may suffer substantial power loss due to the incorporation of a large amount of noises. Overall, existing methods suffer from four main statistical challenges: (C1) they struggle to maintain high statistical power under varying mediation signal structures, such as different levels of signal sparsity, effect size and correlation among the mediators. Some methods are only powerful with more frequent signals, while other methods are only powerful with sparse signals; (C2) the highly correlated nature of mediators is commonly observed in imaging and omics data, and it poses challenges to select and prioritize mediators. Existing methods often select only one or several mediators among a set of highly correlated true mediators; (C3) the dimension reduction procedure in many existing methods leads to violation of causal assumptions (Andrews and Didelez, 2020). (see details in Section 2); (C4) existing methods either miss or have biased estimation of individual mediation contributions due to the natural collinearity among mediators with high exposure effect.

To address the challenges discussed above, we propose a framework with **<u>p</u>**artial **<u>s</u>**um **<u>s</u>**tatistic and **<u>s</u>**ample **<u>s</u>**plitting **<u>s</u>**trategy, namely PS5, for a general high-dimensional causal mediation analysis. Firstly, our method assumes sparsity in mediator-outcome relationship, which means only a small number of mediators would mediate the exposure’s effect on outcome. After that, we apply a sample splitting strategy with penalized regression on the outcome. We prove a proposition to guarantee that the variable selection strategy does not lead to violation of causal assumptions (overcoming C3). By removing marginal exposure effect before penalized regression for variable selection, we successfully avoid biased estimation of individual mediation contributions (overcoming C4). To achieve high statistical power with varying sparsity of mediators, we propose a partial sum statistic to detect the global mediation effect (overcoming C1). By conducting multiple runs of sample splitting, our method further reduces bias in global indirect effect estimation and can successfully identify highly correlated mediators (overcoming C2). Lastly, a series of marginal tests on mediation contribution (Clark-Boucher et al., 2023) can indicate the importance of each mediator, allowing the selection and prioritization of active mediators for insightful biological interpretation. Table 1 summarizes pros and cons of eight existing methods and PS5 based on their capacity to achieve the three aims and to overcome the four statistical challenges.

**Table 1.**
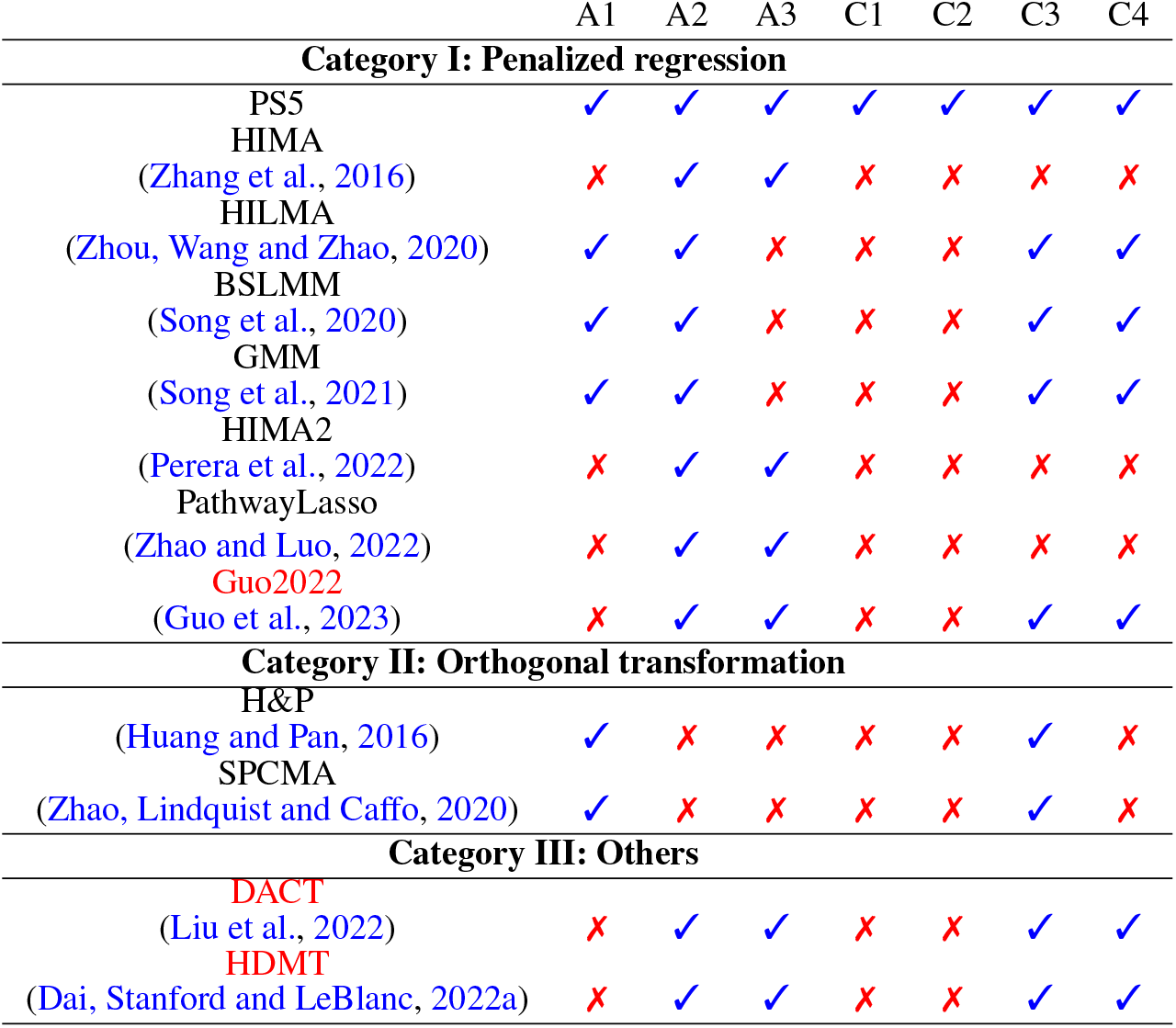
Methods comparison based on three aims and four challenges. A1: Test for global indirect effect; A2: Estimation of global indirect effect; A3: Mediators Prioritization; C1: Capturing unknown signal structure; C2: Feasibility of highly correlated mediators; C3: Rigor of causal assumptions; C4: Unbiased estimation of mediation contributions

We organize this paper as follows. In Section 2, we provide an overview of mediation analysis and causal assumptions needed for identifying causal effects. In Section 3, we introduce PS5 in detail. Section 4 provides extensive simulations to evaluate the performance of different methods based on type I error, power, estimation bias, and sensitivity. In Section 5, we apply PS5 mediation analysis to the imaging genetics dataset in COPDGene using PRS as a genetic exposure, CT imaging as mediators and FEV1 as outcome. We additionally implement a second mediation analysis using smoking (pack-years) as environmental exposure and CT imaging as mediators and integrate the two causal mediation results. Section 6 provides final conclusion and discussion.

## 2. Notations and Assumptions

In this paper, our focus lies on causal mediation analysis involving a group of candidate mediators. Within a high-dimensional setting, suppose we collect a dataset of *N* i.i.d. individuals, denoted as 𝒟 = (*X*_1_, **M**_1_, *Y*_1_, **C**_1_), …, (*X*_*N*_, **M**_*N*_, *Y*_*N*_, **C**_*N*_), where the random variables (*X*, **M**, *Y*, **C**) are generated from a distribution *F*. Each individual has an exposure *X*_*i*_, an outcome *Y*_*i*_, *l*-dimensional covariates 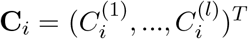, and *p*-dimensional mediators 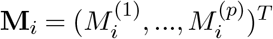, where *i* = 1, …, *N*. To formally define causal mediation effects, we adopt the (counterfactual) potential outcome framework. Specifically, let *Y*_*i*_(*x*, **M**_*i*_(*x*)) represent the potential outcome for subject *i* under the exposure level *x*, and 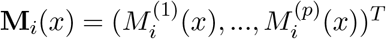 represent a *p*-dimensional potential mediator for subject *i* given the exposure level *x*. We can proceed to decompose the total effect of exposure *X* on outcome *Y* into two components: the direct effect and the effect mediated through the entire group of mediators, known as the indirect effect. The natural direct effect (NDE) is defined as *Y* (*x*, **M**(*x*\*)) − *Y* (*x*\*, **M**(*x*\*)), capturing the effect of *X* on *Y* through pathways that do not involve mediators **M**. On the other hand, the natural indirect effect (NIE) is defined as *Y* (*x*, **M**(*x*)) − *Y* (*x*, **M**(*x*\*)), representing the effect of changing mediators from **M**(*x*\*) to **M**(*x*) when exposure is controlled at level *x*. The total effect (TE) can be decomposed as

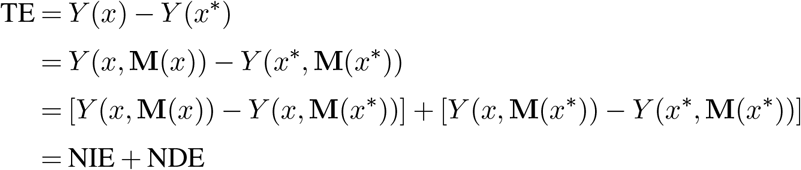

Unlike ideally randomized experiments, which provide a direct way to infer causality, observational studies face greater challenges in establishing causal interpretations due to the absence of randomized treatment assignment (Hernán and Robins, 2020). To establish causal effects, researchers often rely on identification assumptions (Pearl, 2001). VanderWeele and Vansteelandt (2014) propose four identification assumptions for multiple mediators. Denote *A* ⊥⊥ *B*|*C* as *A* independent of *B* conditional on *C*. The following are the sufficient assumptions for identifying the causal effect in mediation analysis:

I. *Y* (*x*) ⊥⊥ *X*|**C**: no unmeasured confounding variables for the exposure-outcome relationship.
II. *Y* (*x*, **m**) ⊥⊥ **M**|*X*, **C**: no unmeasured confounding variables for the mediators-outcome relationship, conditional on the exposure *X*.
III. **M**(*x*) ⊥⊥ *X*|**C**: no unmeasured confounding variables for the exposure-mediators relationship.
IV. *Y* (*x*, **m**) ⊥⊥ **M**(*x*\*)|**C**: no measured or unmeasured *X*-induced confounding variables for the mediators-outcome relationship, conditional on assumption (II).

Assumptions (I) – (III) are the no-unmeasured confounding assumptions, while assumption (IV) is known as the cross-world assumption. It is important to note that when any *X*-induced confounder is present, such as *L* in Figure 1B, assumption (IV) may be violated even if data on *L* is observed (Andrews and Didelez, 2020). In reality, high-dimensional mediators, especially in omics and imaging data, often interact with each other. Consequently, excluding partial mediators from the system can potentially lead to a violation of assumption (IV). For example, in Figure 1C, if *M* ^(1)^ is the cause of *Y* and interacts with at least one of other mediators *M* ^(2)^, …, *M* ^(*p*)^, it becomes the *X*-induced confounder for the multiple mediators model *X*−(*M* ^(2)^, …, *M* ^(*p*)^)−*Y*. Therefore, we should be cautious about dropping mediators from the joint system when reducing the dimensionality of mediators. In section 3.1, we will demonstrate that the proper dimension reduction approach in PS5 can preserve the validity of causal assumptions.

With the above assumptions, the average NDE and NIE can be identified through the following regression models for *Y* and **M** using the observed data:

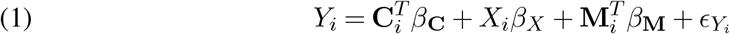

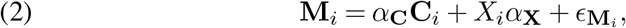

where 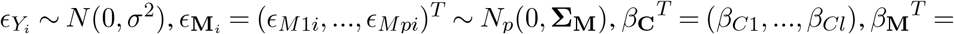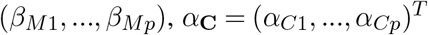 matrix, *α*_**X**_ = (*α*_*X*1_, …, *α*_*Xp*_)^*T*^, and 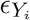 is assumed to be independent with 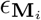.

Here, we assume there is no interaction between *X* and *M*. Then, NDE and NIE can be expressed as below:

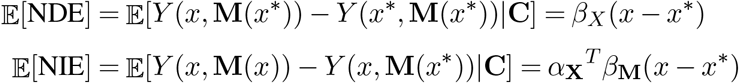

The NDE simply corresponds to the coefficient *β*_*X*_ in the outcome model (1). The NIE can be expressed as the sum of the product of *α*_*Xj*_ and *β*_*Mj*_, *j* = 1, …, *p*. For simplicity in this paper, we will refer to the NDE and NIE as the direct effect and global indirect/mediation effect, respectively. To have a more straightforward interpretation, we also define the 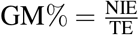 as the global mediation percentage to represent the proportion of the total effect explained by mediators.

## 3. Method

We propose PS5, a three-stage algorithm designed to accomplish three specific aims: (A1) conduct a statistical test for detecting the global indirect effect, (A2) if statistically significant, estimate the global indirect effect, and (A3) select, prioritize and quantify top mediators. Statistically, PS5 addresses four major challenges: (C1) accommodate unknown signal structure with varying sparsity and effect size in mediators, (C2) allow highly correlated mediators, and (C3) preserve causal assumptions (C4) provide unbiased estimation of individual mediation contribution. In the following three subsections, we provide a detailed description of our proposed method.

### 3.1. Sample splitting and variable selection

In this subsection, we illustrate several statistical procedures for variable selection and for ensuring the validity of inferences. Firstly, we adopt a sample splitting procedure to avoid overoptimism of *p*-value assessment resulting from variable selection. Secondly, we remove marginal exposure effects on *Y* and *M* to address high mediator collinearity and instability in the estimation of the mediation effect. Finally, the minimax concave penalty (MCP) method is applied to select candidate mediators for dimension reduction. Proposition 1 is developed to ensure the conservation of causal assumptions (I)-(IV) in the MCP variable selection procedure.

Given the notation in section 2, we assume sparsity in *β*_**M**_ (mediator-outcome relationship), meaning only a small number (*s*; *s* << *N*) of mediators mediate the exposure’s effect on the outcome. In high-dimensional analysis (e.g., *p >> N*), using the data twice for variable selection and statistical inference (i.e., coefficient estimate and *p*-value calculation) can generate overly optimistic results. To this end, we adopt the sample splitting approach (Wasserman and Roeder, 2009; Meinshausen, Meier and Bühlmann, 2009; Dezeure et al., 2015) to circumvent the overfitting. The concept of sample splitting is to divide the data into two equal halves, referred as 𝒟_1_ and 𝒟_2_, each containing *N/*2 observations. In this process, the first-half 𝒟_1_ is utilized to reduce the dimensionality to a manageable size, while the second-half 𝒟_2_ is used for the estimation and *p*-value calculation.

We next propose to remove marginal exposure effect *X* to outcome *Y* and mediators **M** by fitting *Y*_*i*_ = *X*_*i*_*β*_*X*_ + *ϵ*_1_ and **M**_*i*_ = *X*_*i*_*α*_**X**_ + *ϵ*_**2**_, respectively. In simulations and real data, we have found that many mediators are highly correlated due to substantial exposure effect *α*_**X**_, which causes difficulty to estimate *β*_**M**_. Instead of the original *Y*_*i*_ and **M**_*i*_, using 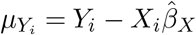 and 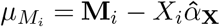 in the MCP variable selection below provides a more accurate estimation of *β*_**M**_, which will be shown in later simulations.

After sample splitting and removing marginal exposure effect, we apply the MCP method proposed by Zhang (2010) to 𝒟_1_:

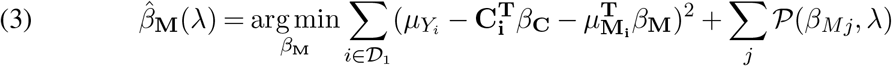

where 𝒫(*β*_*Mj*_, *λ*) is the regularization penalty of MCP. Here we choose MCP due to its less biased estimates for *β*_**M**_ and theoretical consistency in variable selection (Zhang, 2010). Suppose *q* mediators (denoted as *M*′⊂ *M*, *q* << *p*) is selected from 𝒟_1_ by MCP. This selected subset of mediators in 𝒟_2_ will be used in the inferences of the next two subsections.

Proposition 1 below guarantees preservation of causal assumptions (I)-(IV) when we apply the MCP procedure in Equation (3) for mediator selection. Detailed proof is left to Supplementary.

#### Proposition 1

*Given that casual assumptions (I)-(IV) are held for mediators model X*− (*M* ^(1)^, …, *M* ^(*p*)^) −*Y*, *removing candidate mediators without mediator-outcome relationship (as in the MCP procedure in Equation (3)) can still preserve the causal assumptions (I) - (IV)*.

### 3.2. Partial sum statistic for testing global indirect effect

In this subsection, we develop a powerful hypothesis testing procedure for detecting global mediation (indirect) effect using half of the samples that are randomly allocated to 𝒟_2_ with the *q* selected mediators from Equation (3) from 𝒟_1_. A classical hypothesis testing for no global indirect effect is set up as:

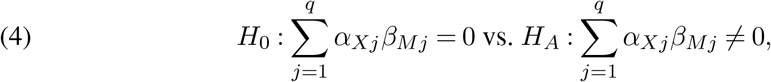

where *α*_*Xj*_*β*_*Mj*_ is also known as the mediation contribution of the *j*^*th*^ mediator. However, as pointed out by Huang and Pan (2016) and Song et al. (2020), the simple sum is less powerful since mediation effect *α*_*Xj*_*β*_*Mj*_ may cancel each other when they have opposite signs. Consequently, we use sum of the *Lγ* norm of *α*_*Xj*_*β*_*Mj*_ for the hypothesis testing set up:

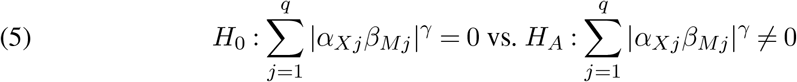

We note that the signals detected by *H*_*A*_ of Equation (4) is a subset of the *H*_*A*_ by Equation (5). For example, a cancellation of effects can happen when the total positive effects of *α*_*Xj*_*β*_*Mj*_ equals the total negative effects, which results in *H*_0_ in Equation (4) but *H*_*A*_ in Equation (5). To better quantify such a cancellation effect, we introduce a measure of neutralization ratio (NR) defined as

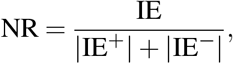

where 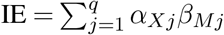 representing global indirect effect, 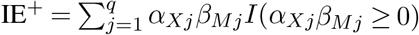, and 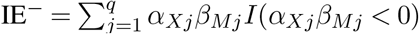.

Below we develop a partial sum statistic for the hypothesis test in Equation (5), which takes into account the sparsity in the exposure-mediator relationship (*α*_**X**_). Mediators without an *α*_**X**_ effect produce zero mediation contribution (*α*_*Xj*_*β*_*Mj*_ = 0) and reduce the statistical power. We first propose the partial sum (PS) score below to allow exclusion of likely null signals to improve power:

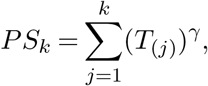

where *T*_*j*_ = |*α*_*Xj*_*β*_*Mj*_|, and *T*_(*j*)_ is the order statistic of *T*_*j*_, and *k* = 1, …, *q*. Denote by *p*_*k*_ the *p*-value of *PS*_*k*_ under the null hypothesis (i.e., 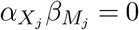 for all 1≤ *j*≤ *q*), of which the detailed calculation will be described in the next paragraph. The final statistic for testing the global mediation effect in Equation (5) is a Cauchy combination test statistic to combine 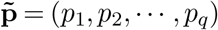:

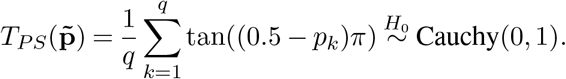

The *p*-value from the global mediation test is then calculated as 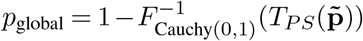. We note that a natural choice for the final combination method could simply by taking the minimum: 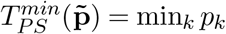 (Li and Tseng, 2011). But since *p*_*k*_’s are dependent, its null distribution has no closed form and requires a second layer of Monte Carlo simulation, making it computationally infeasible in practice. In contrast, the Cauchy combination method has been shown a robust method for combining dependent, sparse and weak signals (Liu and Xie, 2020; Fang, Tseng and Chang, 2023) with null distribution still being a Cauchy distribution. This method is sensitive and robust to detect global signal if any *p*-value in 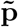 is small.

To calculate *p*_*k*_, we adopt a Monte Carlo method similar to Huang and Pan (2016). Denote by 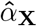 and 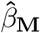 as the maximum likelihood estimator (MLE) of *α*_**X**_ and *β*_**M**_ under the parametric models (1) and (2) using the original data 𝒟 and the second half data 𝒟_2_. Firstly, we approximate the joint distribution of 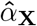 and 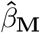 by a multivariate normal distribution,

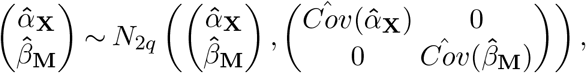

given that *ϵ*_*Y*_ and *ϵ*_**M**_ are independent. Secondly, we generate Monte Carlo samples *α*_**X**_^(*b*)^, *β*_**M**_^(*b*)^ and centered 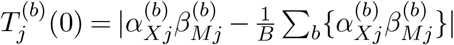, where *b* = 1, …, *B*. Thirdly, we calculate the partial sum statistic for each Monte Carlo sample as 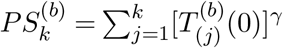 where 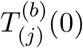 is the order statistic of 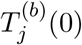. Finally, the *p*-value for *PS*_*k*_ is calculated as 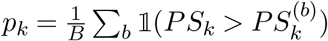 which corresponds to the proportion of Monte Carlo samples where the partial sum statistic is greater than or equal to the observed value.

### 3.3. Multiple sample splitting and prioritization of selected mediators

Although sample splitting provides a valid inference by avoiding over-fitting from variable selection, results from single sample splitting is unstable depending on the sample partition to 𝒟_1_ and 𝒟_2_. Meinshausen, Meier and Bühlmann (2009) proposed multiple sample splitting and a *p*-value aggregation method to avoid obtaining the “*p*-value lottery” result. In our high-dimensional mediator setting, many mediators are correlated. Only one or a few of a set of highly correlated active mediators may be selected in each random sample splitting. To this end, we perform a multiple sample splitting strategy to overcome this issue. The random sample splitting is repeated in parallel for *R* times. Combining results from different 𝒟_1_ in each splitting allows us to capture highly correlated and true mediators. Furthermore, to reduce estimation bias from single sample splitting, we take the median of the estimated global indirect effect from multiple sample splitting iterations. This process helps us achieve more robust and stable results, particularly in cases with highly correlated mediators, by ensuring that our inference is not influenced by the specific data partitioning. In our experience, *R* = 50 is sufficient to generate a stable result while limiting the computational burden.

Based on the mediator sparsity assumption, only a small number of active mediators contribute to the global indirect effect. Prioritizing these key mediators is critical for biological interpretation, decision making, and future investigation. Clark-Boucher et al. (2023) recently pointed out that *α*_*Xj*_*β*_*Mj*_ cannot be directly interpreted as a “causal effect” through the *j*-th mediator. Instead, *α*_*Xj*_*β*_*Mj*_ is named as “mediation contribution” and reflects the active mediation level of the *j*-th mediator, which will be the basis for our prioritization. Following the estimation and inference procedure described in Section 3.2, we can calculate the marginal *p*-value of each mediation contribution *α*_*Xj*_*β*_*Mj*_ using Monte Carlo method:

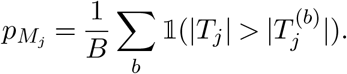

To avoid obtaining the “*p*-value lottery” result, we utilize the *p*-value aggregation method, an empirical *δ*-quantile method suggested by Dezeure et al. (2015), to integrate multiple sample splitting results:

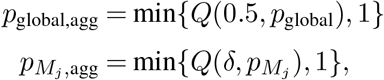

where *Q*(*δ, p*) is the empirical *δ*-quantile of *p*-value vector from *R* multiple sample splitting, and *δ* is set as the half of selected proportion. For example, if mediator *M* ^(1)^ is selected *h* times over *R* multiple sample splitting, *δ* would be set as 0.5*h/R*. The false discovery rate (FDR) and family-wise error rate (FWER) are then further controlled by the Benjamini-Yekutieli and Bonferroni procedures, respectively.

## 4. Simulation

In this section, we compare PS5 with three popular methods reviewed in Zeng, Shao and Zhou (2021), namely H&P (Huang and Pan, 2016), HIMA (Zhang et al., 2016), and HILMA (Zhou, Wang and Zhao, 2020). In our evaluation, we find that H&P tends to have an overestimation issue for 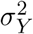 under alternative hypothesis and we apply CV-based LASSO (Reid, Tibshirani and Friedman, 2016) for providing a more robust estimation. Under a moderate sample size (*N*=500), we conduct simulation studies with high-dimensional mediators (*p*=1,000) under different signal strengths and correlation structures. This comprehensive evaluation aims to thoroughly assess the performance of PS5 in aims (A1)-(A3) compared to other applicable methods. For global mediation test in (A1), we evaluate via type I error and statistical power. For estimation of global indirect effect in (A2) and mediators prioritization in (A3), we evaluate the percent of relative bias and sensitivity, respectively. PS5 is generally equal to or more powerful than other methods, has lower estimation bias in mediation effects, and is more accurate in mediator selection.

### 4.1. Type I error of global mediation test

To mimic the PRS exposure, we sample the exposure *X* from *N* (0, 1). The error terms *ϵ*_*Y*_ is generated from *N* (0, 1), and the error terms *ϵ*_**M**_ = (*ϵ*_*M*1_, …, *ϵ*_*Mp*_)^*T*^ are generated *MV N* (0, Σ_*M*_), where Σ_*M* (*a,b*)_ = (*ρ*^|*a*−*b*|^)_*a,b*_ with *ρ* = 0 or 0.5. With *X* and error terms, we generate *p*-dimensional mediators M and outcome Y by using models (1) and (2) with different *α*_**X**_ and *β*_**M**_.

To give a comprehensive comparison, we consider the simulation setting in Dai, Stanford and LeBlanc (2022b) to evaluate type I error under the complete nulls, dense nulls, sparse nulls, and disjunctive nulls. It is important to note that the disjunctive effect sometimes can be interpreted as a mediation effect through the interaction between mediators (Huang and Pan, 2016). However, it is impossible to identify if the causal ordering of mediators is unknown. In this study, we refrain from any causal interpretation of the disjunctive effect and generate four null cases as follows:

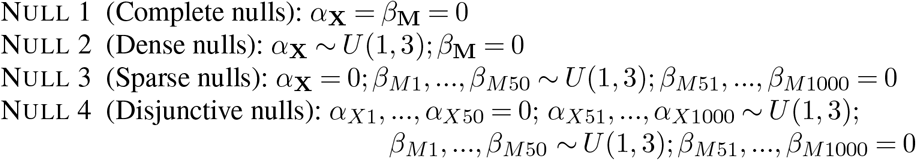

The significance level is set as *p*-value < 0.05. Due to the computational burden of HILMA in some null cases, we only replicate 100 simulations for HILMA, while the other methods are replicated 2,000 times. Table 2 illustrates that only PS5 can reasonably control type I error under 5% across all scenarios. H&P is conservative under complete nulls and sparse nulls and is severely anti-conservative under disjunctive nulls since they treat disjunctive effect as true mediation effect. HIMA is anti-conservative under dense and disjunctive nulls since they perform dimension reduction and make the inference on the same dataset, which is a typical problem in high-dimensional inference. This problem of over-optimism has been discussed by the literature (Meinshausen, Meier and Bühlmann, 2009; Rasines and Young, 2022).

**Table 2.** Type I error results under four null cases and two correlation settings (ρ = 0, 0.5).

|  | Null 1<br>Complete nulls |  | Null 2<br>Dense nulls |  | Null 3<br>Sparse nulls |  | Null 4<br>Disjunctive nulls |  |
| --- | --- | --- | --- | --- | --- | --- | --- | --- |
| | $\rho = 0$ | $\rho = 0.5$ | $\rho = 0$ | $\rho = 0.5$ | $\rho = 0$ | $\rho = 0.5$ | $\rho = 0$ | $\rho = 0.5$ |
| PS5 | 0.00%<sup>†</sup> | 0.00%<sup>†</sup> | 4.50% | 5.45% | 3.10% | 3.75% | 4.85% | 5.65% |
| H&P | 0.00%<sup>†</sup> | 0.00%<sup>†</sup> | 5.15% | 4.47% | 0.00%<sup>†</sup> | 0.00%<sup>†</sup> | 75.05%<sup>‡</sup> | 100%<sup>‡</sup> |
| HIMA | 1.90%<sup>†</sup> | 1.80%<sup>†</sup> | 99.80%<sup>‡</sup> | 99.65%<sup>‡</sup> | 4.75% | 4.35% | 29.65%<sup>‡</sup> | 18.85%<sup>‡</sup> |
| HILMA | 36.00%<sup>‡</sup> | 40.00%<sup>‡</sup> | 100%<sup>‡</sup> | 100%<sup>‡</sup> | 3.00% | 5.00% | 10.00%<sup>‡</sup> | 6.00%<sup>‡</sup> |

<sup>†</sup>: conservative
<sup>‡</sup>: inflated

HILMA is severely anti-conservative when none of *β*_**M**_ exist such as complete and dense nulls. For complete nulls, all methods are either too conservative or overly anti-conservative due to the composite null hypothesis, which remains a challenge in high-dimensional mediation testing.

### 4.2. Power of global mediation test

We design alternative hypotheses with various signal sparsity, signal strengths, and correlation structures, and the power is estimated via 100 simulated data for each setting. Denote by the first *s* candidate mediators (i.e., 𝒮 = 1, …, *s*) as the true mediator set. We assume that the first 50 mediators have *β*_**M**_ effect (*β*_*M*1_ = … = *β*_*M*50_ = 1), while the first *s* true mediators in 𝒮 have *α*_**X**_ effect, where *s* < 50. For signal strengths, we increase *α*_**X**_ magnitude from 0 to 0.2. For correlation structures, we consider one block correlation matrix and two AR1 models, similar to type I error simulations with *ρ* = 0 or 0.5. The block correlation is designed for simulating the true mediators with high correlation. Among the first *s* true mediators, we assume that there are *s/*2 pairs of mediators with correlation=0.9:

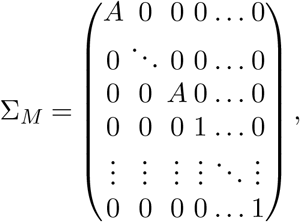

Where 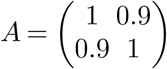. Figure 3A shows the power result of simulations under three correlation settings (*ρ* = 0, 0.5 and 0.9) and three signal structures (|𝒮|/*p* = 0.5%, 1%, 3%). Under sparse scenarios (|𝒮|/*p* = 0.5%), HIMA and PS5 are much more powerful than HILMA and H&P. However, the power of HIMA decreases as correlation increases. When the number of true mediators |𝒮| increases, the power of HIMA becomes lower than PS5 and HILMA (see |𝒮| /*p* = 3%). Overall, PS5 is consistently among the most powerful methods across all scenarios.

**Fig 3:**
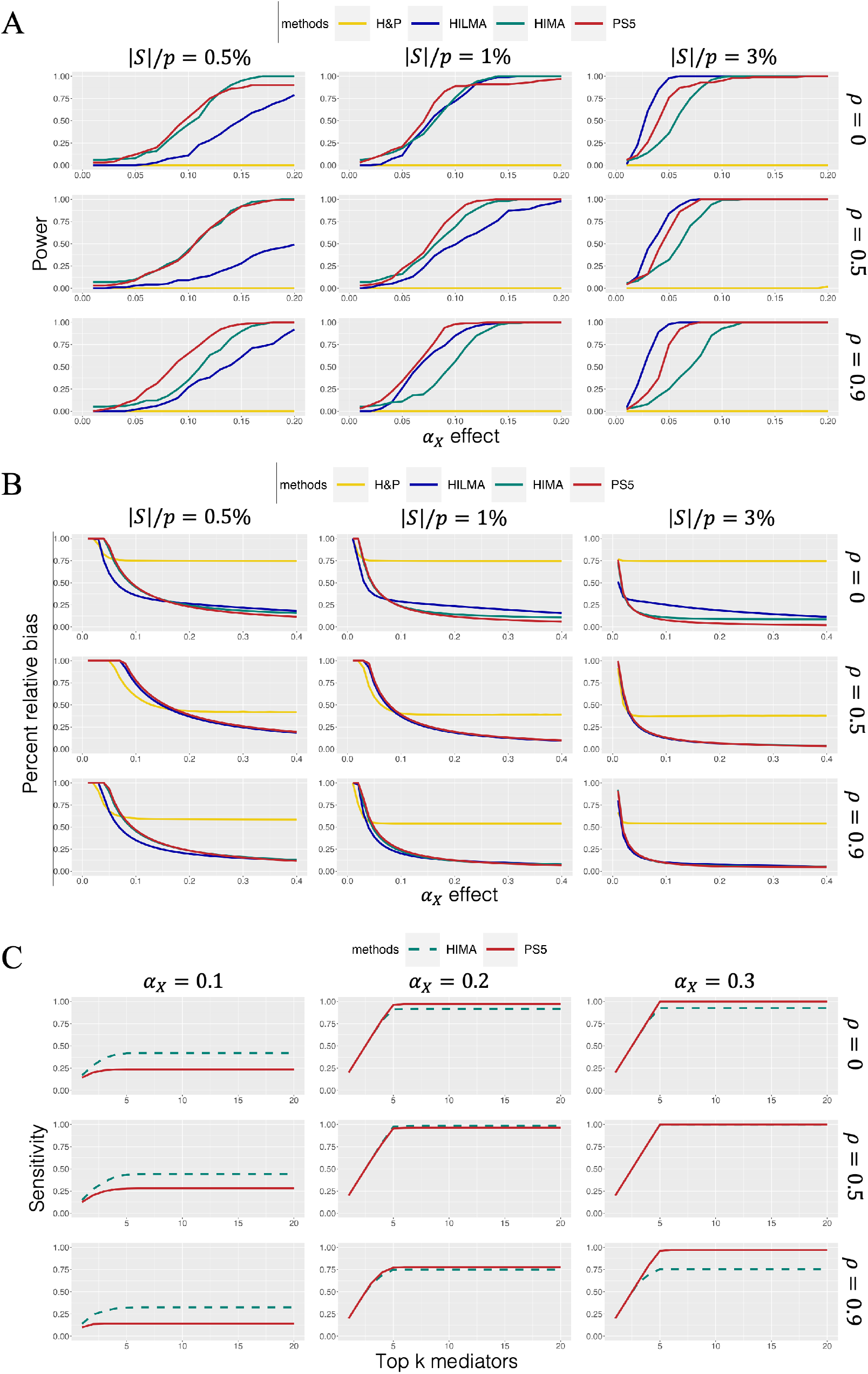
(A) Power for detecting global indirect effect; (B) Percent relative bias for estimating global indirect effect; (C) Sensitivity for mediator prioritization under |𝒮|/*p* = 0.5%.

### 4.3. Estimation of global mediation effect

We assess the relative bias of the global mediation effect, denoted as 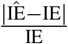 in aim (A2), where IE is the underlying true indirect effect and 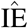 is the estimated indirect effect. Even if H&P method does not provide an unbiased estimation of global mediation effect, we still include H&P for a comprehensive comparison. In Figure 3B, we show relative bias of four methods under three correlation structures and three signal structures, which is the same setting as in Figure 3A for power comparison. However, we increase signal strengths *α*_**X**_ from 0 to 0.4 for visualizing the stable estimation bias. Under the non-correlation setting (*ρ* = 0), HILMA is roughly 10% higher than HIMA and PS5. In the other settings with correlated mediators, PS5, HIMA, and HILMA can achieve lower estimation bias. Among these four methods, H&P tends to have much higher relative bias, regardless of the correlation.

### 4.4. Mediator prioritization

We next evaluate the accuracy of mediator prioritization by sensitivity (i.e., the proportion of true positives among all true mediators) when the top *k* mediators are claimed. Since HILMA and H&P do not provide *p*-value for individual mediators, we only compare PS5 and HIMA. Figure 3C shows the sensitivity results under |𝒮|/*p* = 0.5%, and the other two scenarios (|𝒮|/*p* = 1% and 3%) are shown in Supplementary. In the weaker *α*_**X**_ magnitude, HIMA has slightly higher sensitivity because PS5 makes inferences based on half data (𝒟_2_). However, when *α*_**X**_ magnitude increases, only PS5 can eventually reach 100% sensitivity, especially in block correlation design with high correlation. On the other hand, HIMA cannot select all true mediators even if the signal strength is strong. The reason is that HIMA applies penalized regression, which has the limitation of accurately selecting highly correlated variables. The result shows that multiple sample splitting in Section 3.3 in PS5 successfully overcomes the issue of detecting highly correlated mediators.

## 5. Imaging genetics application in the COPDGene study

Chronic obstructive pulmonary disease (COPD) is ranked as the third leading cause of mortality worldwide, accounting for 3 million deaths in 2019 alone (Mei et al., 2022). While multiple environmental and social factors, such as cigarette smoking, are associated with an individual’s susceptibility to developing COPD (Salvi, 2014), it is also recognized as a heterogeneous disease (Regan et al., 2011). Existing genome-wide association studies have reported many single nucleotide polymorphisms (SNPs) as potential genetic risk factors for COPD although the effect of each individual SNP is typically small (Pillai et al., 2009; Cho et al., 2014; Lutz et al., 2015; Siedlinski et al., 2013).

COPDGene is a large consortium study with complete genetics and CT imaging information (N=8,897), aiming to investigate the underlying genetic factors of COPD (Regan et al., 2011). In our mediation applications below, CT images will serve as potential mediators. The 3D pixel-resolution images with 512 pixels×512 pixels per slice and more than 512 slices are pre-processed by a self-supervised representation learning method (Li, Ke and Kayhan, 2021), which generates 128 representations for each of the 581 patches (local regions) (i.e. 128 × 581 = 74, 368 features). We then apply principal component analysis (PCA) to each patch and select the first 10 principal components (∼ 80% explained variance) as our final candidate mediators, resulting in *p* = 5, 810 candidate mediators. Each principal component is labeled as “patch index - PC”. For example, M90-1 in Table 3 represents the first PC of the 90-th patch. For the outcome variable, we use forced expiratory volume in one second (FEV1), the amount of air a person can force out from lung in one second, as a surrogate of the disease severity. Since each individual SNP has a small genetic effect on outcome, we employ a polygenic risk score (PRS), which aggregates an individual’s genetic risks from selected SNPs, as the genetic exposure variable in the first mediation application (Moll et al., 2020). In contrast to the genetic factor, cigarette smoke is recognized as the most important causative factor (Laniado-Laborín, 2009) since smoke-induced damage to lung or airway wellness can exacerbate the progression of COPD. Consequently, we perform a second mediation analysis using cigarette smoke as an environmental exposure, which is quantified in pack-years (PY). We include three commonly used covariates in COPD research (i.e., sex, height, and age) in both mediation analyses. We note that genetic exposure (PRS) and environmental exposure (PY) have no correlation (*ρ* = 0.00059) as expected.

**Table 3.** Top mediators for PRS and Pack Years exposures ordered by Fisher’s Combined p-value.

|  | *p*-value |  | Med contri |  | Contri prop |  | Fisher's<br>Combined<br>*p*-value |
| --- | --- | --- | --- | --- | --- | --- | --- |
|  | PRS | Pack Years | PRS | Pack Years | PRS | Pack Years |  |
| M90-1 | **2.49e-04\*\*** | **2.24e-04\*\*** | -0.2856 | -0.1728 | 4.94% | 4.78% | **9.90e-07\*\*** |
| M142-1 | **2.79e-04\*\*** | **2.02e-04\*\*** | -0.1504 | -0.1216 | 2.60% | 3.38% | **1.00e-06\*\*** |
| M233-1 | **3.43e-04\*\*** | **2.59e-04\*\*** | -0.1361 | -0.1120 | 2.35% | 3.10% | **1.53e-06\*\*** |
| M451-1 | **3.44e-04\*\*** | **6.82e-04\*\*** | -0.2423 | -0.1280 | 4.19% | 3.52% | **3.82e-06\*\*** |
| M133-3 | **2.12e-03\*** | **2.58e-04\*\*** | -0.1156 | -0.1536 | 2.00% | 4.25% | **8.46e-06\*\*** |
| M452-1 | **1.86e-03\*** | **1.33e-03\*** | -0.2332 | -0.1104 | 4.03% | 3.08% | **3.47e-05\*\*** |
| M68-1 | **4.36e-03\*** | **1.76e-03\*** | -0.1640 | -0.1184 | 2.83% | 3.27% | **9.85e-05\*\*** |
| M148-1 | **2.16e-03\*** | **3.64e-03\*** | -0.2188 | -0.2112 | 3.78% | 5.81% | **1.00e-04\*\*** |
| M428-1 | **2.08e-03\*** | 1.24e-02 | -0.1230 | -0.0592 | 2.12% | 1.64% | **3.01e-04\*\*** |
| M445-3 | **6.63e-03\*** | **4.77e-03\*** | -0.1040 | -0.0688 | 1.79% | 1.91% | **3.59e-04\*\*** |
| M60-2 | 1.41e-02 | **6.95e-03\*** | -0.0912 | -0.0624 | 1.57% | 1.73% | **1.00e-03\*** |
| M133-1 | 4.78e-02 | **2.25e-03\*** | -0.0915 | -0.1024 | 1.58% | 2.83% | **1.09e-03\*** |
| M525-2 | 1.39e-02 | **8.58e-03\*** | -0.0978 | -0.0784 | 1.69% | 2.16% | **1.20e-03\*** |
| M149-2 | **9.73e-03\*** | 1.45e-02 | -0.1945 | -0.0928 | 3.36% | 2.56% | **1.39e-03\*** |
| M166-2 | 4.96e-02 | **8.53e-03\*** | -0.0727 | -0.0640 | 1.25% | 2.04% | **3.71e-03\*** |
| M493-8 | 1.18e-01 | **4.32e-03\*** | -0.0286 | -0.0464 | 0.49% | 1.29% | **4.39e-03\*** |
| M545-1 | 1.00e-00 | **7.56e-04\*\*** | -0.0133 | -0.1328 | 0.23% | 3.67% | **6.19e-03\*** |
| M70-1 | **4.45e-03\*** | 2.20e-01 | -0.1880 | -0.0864 | 3.25% | 2.37% | **7.78e-03\*** |
| M132-4 | 1.93e-01 | **5.83e-03\*** | -0.0361 | 0.0528 | 0.62% | 1.46% | **8.79e-03\*** |
| M579-1 | 8.57e-01 | **2.01e-03\*** | 0.0396 | -0.0896 | 0.68% | 2.48% | 1.26e-02 |
| M207-1 | 6.30e-01 | **3.02e-03\*** | -0.0744 | -0.1600 | 1.28% | 4.42% | 1.38e-02 |
| M303-1 | 1.00e-00 | **2.57e-03\*** | -0.0198 | -0.0896 | 0.34% | 2.47% | 1.79e-02 |
| M141-2 | **2.81e-03\*** | 1.00e-00 | -0.1346 | -0.0160 | 2.33% | 0.48% | 1.93e-02 |
| M553-2 | 1.00e-00 | **3.29e-03\*** | -0.0306 | -0.1072 | 0.52% | 2.98% | 2.21e-02 |
| M96-2 | 1.00e-00 | **5.57e-03\*** | -0.0146 | -0.0864 | 0.25% | 2.40% | 3.44e-02 |

"\*" denotes *p*-value < 0.01; "\*\*" denotes *p*-value < 0.001
"Med contri" denotes the mediation contribution of a single mediator.
"Contri prop" denotes the proportion of mediation contribution (total effect/mediation contribution).
"Fisher's Combined *p*-value" denotes the combined *p*-value by Fisher's method.

In the first mediation analysis using PRS as the genetic exposure, we pursue the three aims (A1)-(A3) using the PS5 method: (1) A global mediation testing to decide whether CT imaging is a mediator as a whole in the impact of PRS on FEV1; (2) If CT imaging is a statistically significant mediator, we estimate its global mediation percentage (GM%) as the proportion of total effect mediated by imaging; (3) Prioritize the top lung patches and quantify the mediation contribution in each patch. For aim 1, the result identifies CT imaging as a strong mediator between PRS and FEV1 with a significant *p*-value (*p* < 10^−16^) and low neutralization rate (21%). The result indicates that global indirect effect exists and the marginal mediation contribution from different active patches mostly present the same sign (direction) of mediation contribution. In aim 2, we estimate 49% of the total effect between PRS and FEV1 is mediated through lung image (i.e. GM% = 49%). We then conclude that CT image as a pulmonary wellness surrogate has a strong mediation effect between PRS and FEV1, which could have potential clinical implications if a treatment or intervention can target the active mediation regions. For this purpose, we identify 13 significant patches with significant mediation contribution (*p* < 0.01) in aim 3. Table 3 shows the patch IDs, *p*-values, mediation contributions, and contribution proportions of the 13 significant patches (marked with “*” or “**” in the second column reflecting *p* < 0.01 or *p* < 0.001). The contribution proportion, here, is defined as the percentage of total effect.

We next perform a second mediation analysis using smoking pack-years (PY) as the environmental exposure and similarly pursue the three aims. The result of aim 1 also shows that CT imaging as a whole is a significant mediator in the impact of smoking on FEV1 (*p* < 10^−16^). The low neutralization rate (18%) also similarly shows low cancellation of positive and negative mediation effects among individual active mediators. Aim 2 shows that 76% of the total effect between smoke and FEV1 is mediated through CT imaging (i.e. GM% = 76%), a magnitude higher than 49% in the PRS mediation analysis. In aim 3, Table 3 shows 20 significant patches detected under *p* < 0.01. Similarly, the 20 significant patches in this environmental mediation are highlighted in the third column with “*” or “**” reflecting *p* < 0.01 or *p* < 0.001.

Since PRS and PY have almost zero correlation and represent genetic and environmental exposures, it is of interest to see whether the two mediation analyses identify similar patches in CT imaging. If so, these overlapped physical regions in lung might suggest disease-related mechanisms (e.g., inflammation or immune response) in certain enriched tissues or cell types, which may lead to targeted therapy or intervention to slow the disease progression. To this end, we include all 25 mediators (24 patches) in Table 3 as the union set of the 13 patches detected by PRS-induced mediation analysis and 20 patches detected by PY-induced mediation analysis, where the 25 mediators are ordered by the meta-analyzed *p*-values using Fisher’s method (the last column). Of the 9 patches overlapped by the 13 and the 20 detected patches under *p* < 0.01, the 2 × 2 table in Figure 4A shows an overlap enrichment of *p*-value = 5.7 × 10^−12^ from Fisher’s exact test and odds ratio= 108.26. To visualize the 3D locations of detected patches, Figure 4B shows histograms of the marginal counts of detected patches in varying *X, Y*, and *Z* coordinates (red: detected by both, blue: PRS only, and orange: PY only). It is worth noting that *X*-coordinate shows two clusters representing right and left lungs. *Z*-coordinate shows active regions mostly in the lower lobe; particularly all PRS and PY overlapped patches are in the lower lobe (*Z* = 108 ∼ 160). Figure 4C shows eight slices of 2D images at varying *Z*-coordinates. Notably, a cluster of four active mediation patches (M90, M148, M133, and M68) overlapped from PRS-induced and PY-induced mediation analysis at *Z* = 108. Note that some patches in Figure 4C lie outside the lung region (e.g., M207, M233, M303, and M428) since we use one subject for visualization. Our feature extraction method selects patches based on the average frequency of the patch that lies inside the lung across the population. In other words, a patch is considered when, in the majority of cases, the patch is inside the lung.

**Fig 4:**
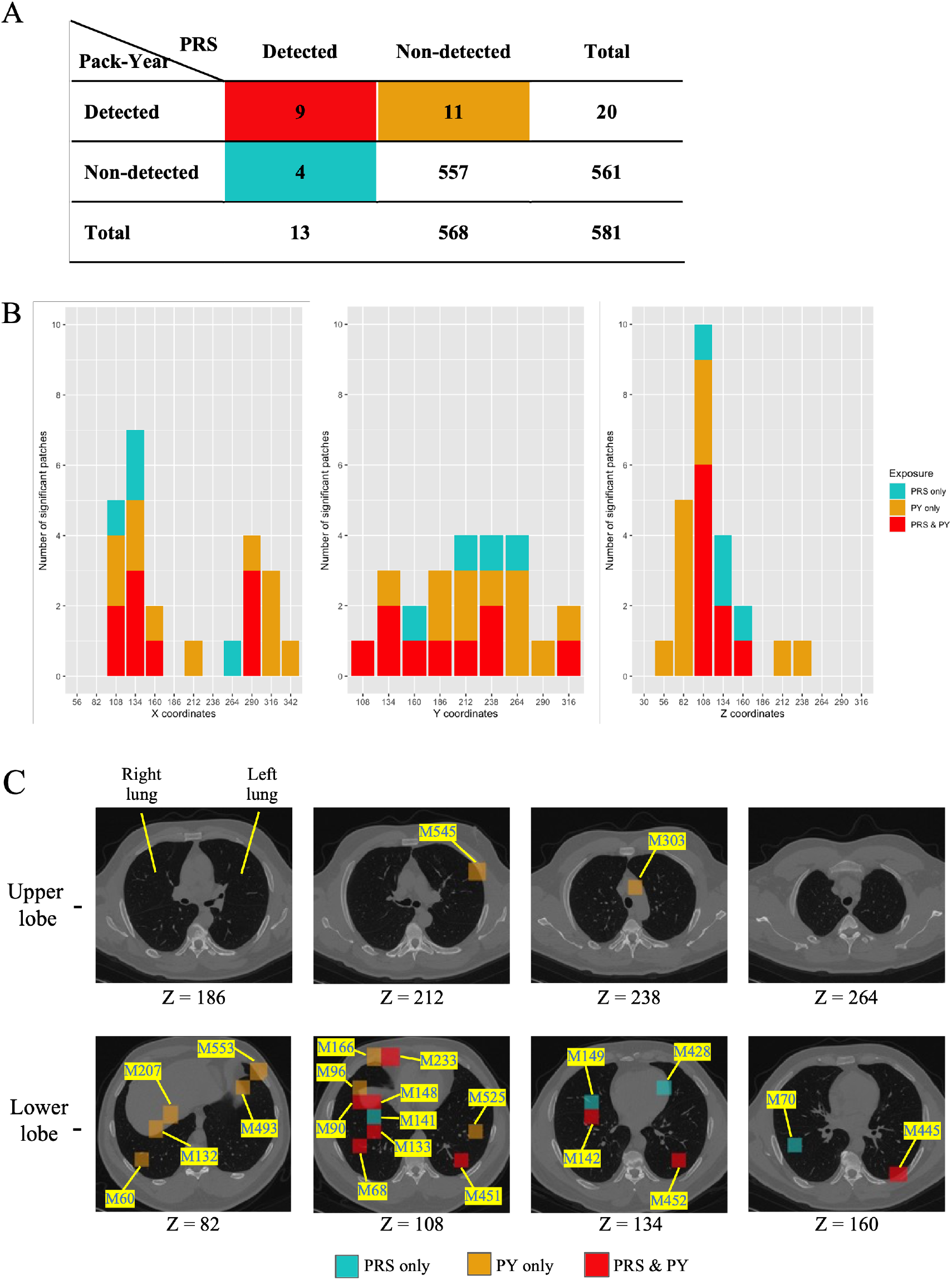
Visualization of COPD Mediation Analysis. (A) 2×2 contingency table of detected mediators from PRS-induced and PY-induced mediation analysis. (B) Histogram of Significant Patches: The histogram displays the distribution of significant patches along the *X, Y*, and *Z* coordinates of the lung image. (C) CT Images on Different *Z*-Coordinates: These images visualize the most significant patches located in the lower lobe.

We identify lower lobes as the significant CT imaging regions, which has strong mediation effects for both smoke as an environmental exposure and PRS as a genetic exposure (Figure 4C). Unlike other well-studied organ imaging related to diseases such as neuroimaging for psychiatric disorders, there is little understanding of lung imaging related to diseases such as COPD and asthma. Limited existing studies have shown that COPD has a greater impact on the upper lobes (Takahashi et al., 2008), which is reasonable since upper lobes are closer to smoke inhaling. However, our finding of right lower lobe as the focused and overlapped mediation region from both mediation analyses using genetic or environmental exposure is surprising and could be clinically impactful. The paradigm shifting understanding in CT imaging may offer possibilities of progression prediction, targeted treatment, or intervention towards the mediated subregions.

## 6. Discussion

Causal mediation analysis has provided an impactful role in observational studies to infer the causal roles of mediators through which an exposure influences the outcome. With availability of high-dimensional mediators using omics or imaging studies, the need of a powerful and accurate framework for high-dimensional causal mediation is emerging. Our proposed PS5 aims to answer three sequential questions in high-dimensional causal mediation (A1-A3): firstly whether the global mediation effect is statistically significant, secondly the proportion of association effect is through the set of mediators, and finally identification of active mediators and a ranked list of their contribution. Multiple innovative statistical procedures, such as partial sum statistic and (multiple) sample splitting, are employed to overcome four statistical challenges (C1-C4), including achieving high statistical power under different mediation signal structures, accurately detecting highly correlated true mediators, ensuring causal assumptions, and accurate estimation of individual mediation contributions. Extensive simulations and a real application in COPDGene imaging genetics mediation analyses both show superior performance and insightful biological findings of PS5, compared to existing methods.

The choice of *γ* in Equation (5) has an impact of statistical power on detecting frequent or sparse signal. According to the simulation of the *γ* parameter (see Supplementary), *γ* = 2 provides higher power than *γ* = 1 for detecting sparse signal. A larger *γ* increases the influence of the one or several strongest signals and is thus more powerful for sparse signal, a situation similar to using heavy-tailed distribution transformation for combining *p*-values discussed in Fang, Tseng and Chang (2023). Therefore, we recommend using *γ* = 2 for providing a good trade-off to achieve high statistical power in varying levels of signal sparsity.

Although PS5 involves sample splitting and Monte Carlo procedures, the computational burden, in terms of speed and memory demand, is reasonable. In the COPDGene application using PRS as exposure, we have *N*= 8, 897 patients and *p* = 5, 810 candidate mediators. Using a Dell server with 32 cores (Intel Xeon Gold 5218) and 128GB RAM, the analysis of 500 multiple sample splitting requires 9.33 minutes (on 64 threads), compared to 2.68 minutes for HIMA, 18.83 minutes for H&P, and 4.16 hours for HILMA. Since multiple sample splitting can easily be implemented in parallel, GPU and parallel computing can easily be incorporated to further reduce computing time.

We note that PS5 is a general framework for high-dimensional candidate mediators. The current PS5 does not consider the spatial structure and potential correlation of mediation effects among patches in CT imaging, which is a future direction. An R software package is available at https://github.com/hung-ching-chang/PS5Med with data and code included for reproducing all results in this paper.

## Data Availability

The data is not publicly accessible as it is part of NIH-sponsored clinical trials and requires a signed data-use agreement. For access to COPDGene data, please visit https://www.copdgene.org for instructions.

## Acknowledgments

We are grateful to the editor, the associate editor, and the referees for their helpful comments. We thank Dr. Yen-Tsung Huang for the insightful discussions.

## SUPPLEMENTARY MATERIAL

### Supplementary Materials for

The online supplemental materials include proof of Proposition 1, comparison of *γ* parameter in (5), two additional sensitivity results for continuous exposure (|𝒮|/*p* = 1% and 3%), and simulation results for discrete exposure.

