## Supplementary Materials for "High-dimensional causal mediation analysis by partial sum statistic and sample splitting strategy in imaging genetics application"

### Supplementary Materials for "High-Dimensional Causal Mediation Analysis by Partial Sum Statistic and Sample Splitting Strategy in Imaging Genetics Study"

Hung-Ching Chang, Yusi Fang, Michael T. Gorczyca, Kayhan Batmanghelich,

George C. Tseng*

#### S1 Proof of Proposition 1

*Proof.* Denote $X$ as exposure, $Y$ as outcome, and $(M^{(1)}, ..., M^{(p)})$ as a $p$-dimensional mediator. Given the casual assumptions (I)-(IV) hold for multiple mediators model $X-$ $(M^{(1)}, ..., M^{(p)})$ $-Y$, we consider the corresponding Pearl's non-parametric structural equation model (Pearl, 2001) with a p-dimensional mediator:

$$X = f_X(\epsilon_X)$$
$$\mathbf{M} = f_{\mathbf{M}}(X, \epsilon_{\mathbf{M}})$$
$$Y = f_Y(X, \mathbf{M}, \epsilon_Y),$$

where $\mathbf{M} = (M^{(1)}, ..., M^{(p)})^T$, and the $\epsilon$ are noise terms. Here, for simplicity, we ignore covariates $\mathbf{C}$, but the proof can be extended with a suitable set of $\mathbf{C}$. With general setting of mediation model, noise terms are jointly independent ($\epsilon_X \perp\!\!\!\perp \epsilon_{\mathbf{M}}, \epsilon_X \perp\!\!\!\perp \epsilon_Y, \epsilon_{\mathbf{M}} \perp\!\!\!\perp \epsilon_Y$) (Peters et al., 2017).

*Correspond to:

Suppose that the $M^{(j)}$ is not the cause of the outcome, which means $M^{(j)}{-}Y$ relationship does not exist, i.e.,

$$Y = f_Y(X, \mathbf{M}^{(-j)}, \epsilon_Y).$$

We demonstrate the proof by considering $\mathbf{M}^{(-j)} = \{\mathbf{M}_{in}^{(-j)}, \mathbf{M}_{out}^{(-j)}, \mathbf{M}_{no}^{(-j)}\}$, where $\mathbf{M}_{in}^{(-j)}$, $\mathbf{M}_{out}^{(-j)}$, and $\mathbf{M}_{no}^{(-j)}$ are the three distinct subsets: $\mathbf{M}_{in}^{(-j)}$ is the cause of $M^{(j)}$, $\mathbf{M}_{out}^{(-j)}$ is affected by $M^{(j)}$, and $\mathbf{M}_{no}^{(-j)}$ does not have direct interaction with $M^{(j)}$ (Figure S1). More general, $\mathbf{M}_{in}^{(-j)}$, $\mathbf{M}_{out}^{(-j)}$, and $\mathbf{M}_{no}^{(-j)}$ may interact with each other. Based on these, the model $\mathbf{M}$ can be divided into $M^{(j)}$ model, $\mathbf{M}_{in}^{(-j)}$ model, $\mathbf{M}_{out}^{(-j)}$ model, and $\mathbf{M}_{no}^{(-j)}$ model, i.e.,

$$\begin{aligned}
M^{(j)} &= f_{M^{(j)}}(X, \mathbf{M}_{in}^{(-j)}, \epsilon_{M^{(j)}}) \\
\mathbf{M}_{in}^{(-j)} &= f_{\mathbf{M}_{in}^{(-j)}}(X, \mathbf{M}_{out}^{(-j)}, \mathbf{M}_{no}^{(-j)}, \epsilon_{\mathbf{M}_{in}^{(-j)}}) \\
\mathbf{M}_{out}^{(-j)} &= f_{\mathbf{M}_{out}^{(-j)}}(X, M^{(j)}, \mathbf{M}_{in}^{(-j)}, \mathbf{M}_{no}^{(-j)}, \epsilon_{\mathbf{M}_{out}^{(-j)}}) \\
\mathbf{M}_{no}^{(-j)} &= f_{\mathbf{M}_{no}^{(-j)}}(X, \mathbf{M}_{in}^{(-j)}, \mathbf{M}_{out}^{(-j)}, \epsilon_{\mathbf{M}_{no}^{(-j)}}).
\end{aligned}$$

To simplify the result, we combine three $\mathbf{M}^{(-j)}$ models with considering sufficient input $X$, $M^{(j)}$, and $\epsilon_{\mathbf{M}^{(-j)}}$,

$$\mathbf{M}^{(-j)} = f_{\mathbf{M}^{(-j)}}(X, M^{(j)}, \epsilon_{\mathbf{M}^{(-j)}}),$$

where $\epsilon_{M^{(j)}} \perp\!\!\!\perp \epsilon_{\mathbf{M}^{(-j)}}$, $\epsilon_{M^{(j)}} \perp\!\!\!\perp \epsilon_X$, $\epsilon_{M^{(j)}} \perp\!\!\!\perp \epsilon_Y$, $\epsilon_{\mathbf{M}^{(-j)}} \perp\!\!\!\perp \epsilon_X$, and $\epsilon_{\mathbf{M}^{(-j)}} \perp\!\!\!\perp \epsilon_Y$. This system of equations implies the following potential outcome model:

$$\begin{aligned}
M^{(j)}(x) &= f_{M^{(j)}}(x, \mathbf{M}_{in}^{(-j)}(x), \epsilon_{M^{(j)}}) \\
M^{(j)}(x^*) &= f_{M^{(j)}}(x^*, \mathbf{M}_{in}^{(-j)}(x^*), \epsilon_{M^{(j)}}) \\
\mathbf{M}^{(-j)}(x^*) &= f_{\mathbf{M}^{(-j)}}(x^*, M^{(j)}(x^*), \epsilon_{\mathbf{M}^{(-j)}}) \\
Y(x, \mathbf{m}^{(-j)}) &= f_Y(x, \mathbf{m}^{(-j)}, \epsilon_Y)
\end{aligned}$$

This implies that assumption (IV) hold, $Y(x, \mathbf{m}^{(-j)}) \perp\!\!\!\perp \mathbf{M}^{(-j)}(x^*)$, since they do not share the same structure of the joint distribution.

In summary, if $M^{(j)}$ is not a parent of $Y$, then it appears trivial that the four assumptions hold under Rubin's potential outcomes framework. Particularly if these four assumptions are made prior to removal of $M^{(j)}$. Assumption (I) doesn't concern the mediator-outcome relationship. Assumption (II) should hold by consistency, as $M^{(j)}$ would not become confounding variables for the mediator-outcome relationship. Assumption (III) has nothing to do with the mediator-outcome relationship. Assumption (IV) should again hold by consistency. □

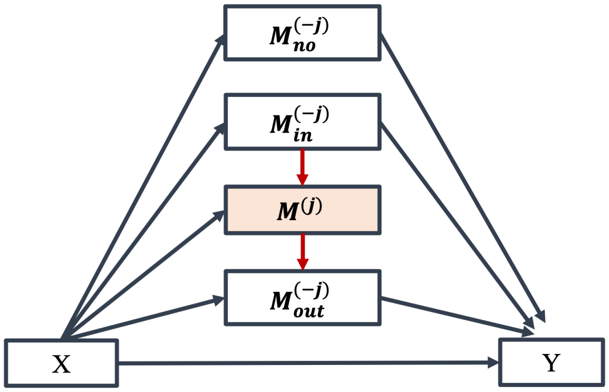


Figure S1: Example of a directed acyclic graph representing mediation effect through a set of mediators $\mathbf{M}$, but there is a candidate mediator $M^{(j)}$ without a mediator-outcome relationship.

### S2 Comparison of $\gamma$ parameter

We consider two $\gamma$ parameters ($\gamma = 1$ and 2), which represent the $L1$ and $L2$ norm of $\alpha_{Xj}\beta_{Mj}$ in null hypothesis and partial sum statistic, under non-correlation setting ($\rho = 0$) and four signal structures ($|\mathcal{S}|/p = 0.5\%$, 1%, 3%, and 5%). Figure S2 shows that $\gamma = 2$ provides roughly 10% higher power than $\gamma = 1$ for detecting sparse signal ($|\mathcal{S}|/p = 0.5\%$ and 1%), while the power of two $\gamma$ settings are similar for detecting non-sparse signal ($|\mathcal{S}|/p = 3\%$ and 5%). A larger $\gamma$ increases the influence of the one or several strongest signals and is thus more powerful for sparse signal.

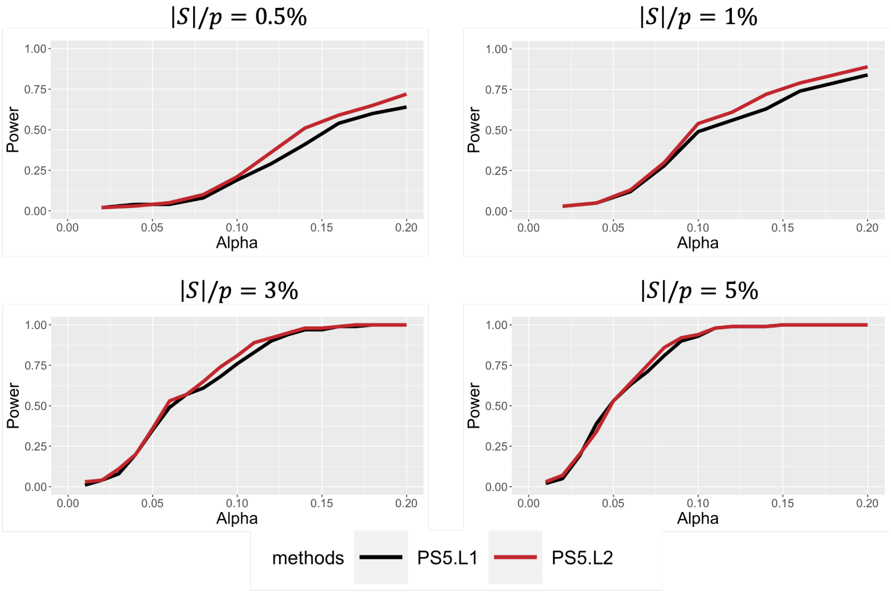


Figure S2: Power of two $\gamma$ settings for detecting global indirect effect under four signal structures ($|\mathcal{S}|/p = 0.5\%$, 1%, 3%, and 5%).

#### S3 Additional sensitivity results for $|\mathcal{S}|/p = 1\%$ and $3\%$

We provide two additional sensitivity results for different signal structures ($|\mathcal{S}|/p = 1\%$ and 3%) in Figure S3. Regardless of signal structures, PS5 can reach 100% sensitivity as $\alpha_{\mathbf{X}}$ magnitude increases. However, HIMA cannot select all true mediators even if the signal strength is strong.

#### S4 Simulation results for discrete exposure

To mimic the SNPs exposure, we also do the comprehensive simulation by randomly sampling discrete exposure $X$ from 0, 1, and 2. All other parameter settings remained consistent with those in the main article. We present results for type I error, power, relative bias, and accuracy of mediator prioritization in Table S1, Figure S4, and Figure S5.

For type I error control (Table S1), only PS5 successfully controls type I error under 5% across all scenarios, while other methods tend to be inflated or conservative. Under Null 1, all methods have conservative type I error. For the power of global mediation test (Figure S4A), HIMA and PS5 have higher power than HILMA and H&P. However, as the number of true signals $|\mathcal{S}|$ increases, HILMA and PS5 become the most powerful methods across all four methods. Overall, PS5 was the only method capable of effectively detecting both sparse and non-sparse signals.

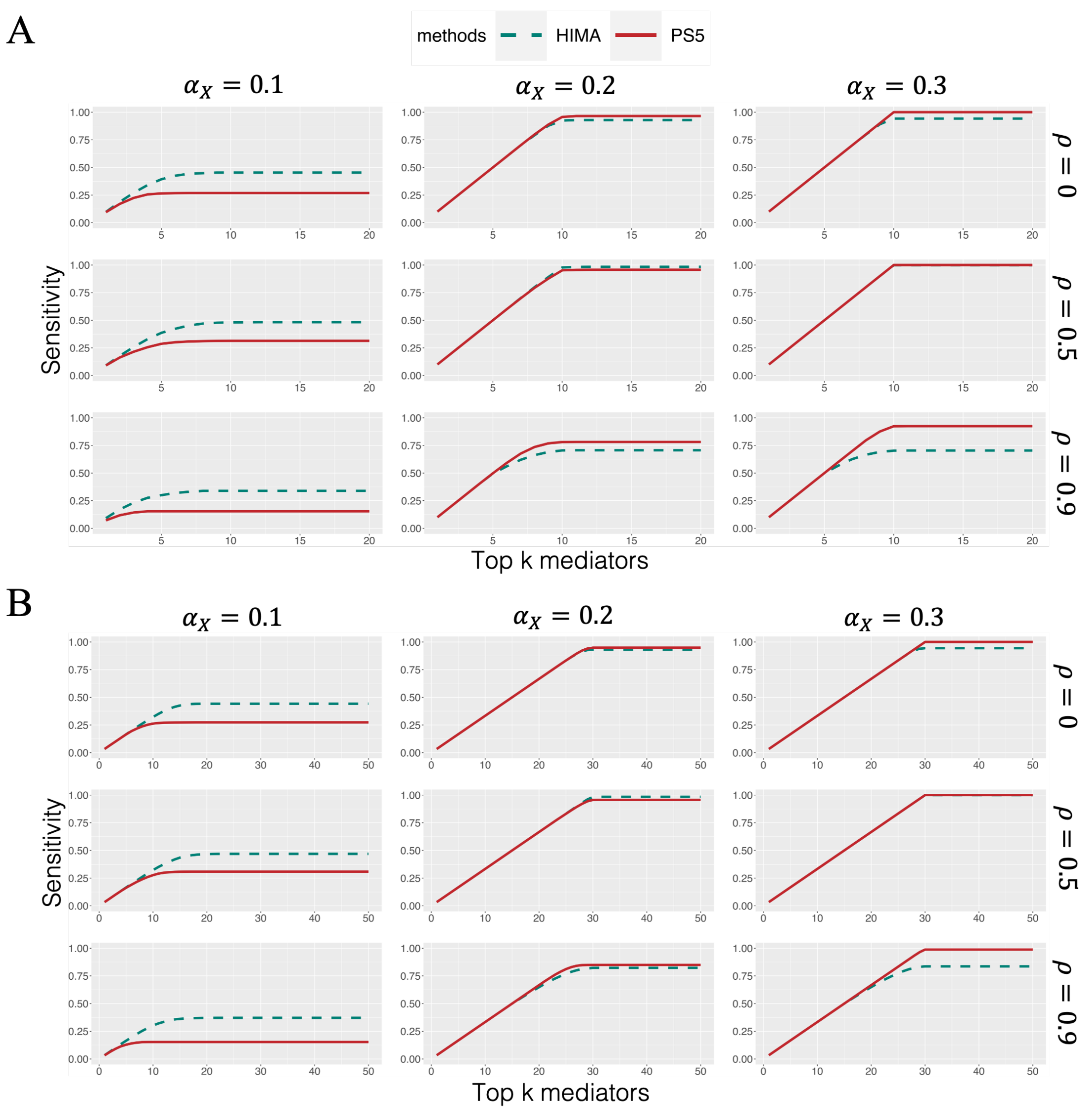


Figure S3: Accuracy of mediator prioritization for continuous exposure by sensitivity. (A) $|\mathcal{S}|/p = 1\%$ (B) $|\mathcal{S}|/p = 3\%$

For relative estimation bias of the global mediation effect (Figure S4B), PS5 would be the best-performing method and roughly $10 \sim 20\%$ lower than HILMA and HIMA under non-correlation settings. In the other settings with correlated mediators, both PS5 and HILMA can achieve lower estimation bias, which outperforms HIMA by approximately 10%. Among these three methods, the relative bias of H&P tends to be much higher, regardless of the correlation structures. For the accuracy of mediator prioritization, we present the result by sensitivity, and Figure S5 shows that only PS5 can achieve 100% sensitivity when $\alpha_{\mathbf{X}}$ magnitude increases.

| | Null 1<br>Complete nulls | | Null 2<br>Dense nulls | | Null 3<br>Sparse nulls | | Null 4<br>Disjunctive nulls | |
|---|---|---|---|---|---|---|---|---|
| | $\rho = 0$ | $\rho = 0.5$ | $\rho = 0$ | $\rho = 0.5$ | $\rho = 0$ | $\rho = 0.5$ | $\rho = 0$ | $\rho = 0.5$ |
| PS5 | 0.00%$^{\dagger}$ | 0.00%$^{\dagger}$ | 5.60% | 5.90% | 3.25% | 3.95% | 5.60% | 5.30% |
| H&P | 0.00%$^{\dagger}$ | 0.00%$^{\dagger}$ | 5.10% | 5.10% | 0.00%$^{\dagger}$ | 1.20%$^{\dagger}$ | 100%$^{\ddagger}$ | 100%$^{\ddagger}$ |
| HIMA | 1.80%$^{\dagger}$ | 1.55%$^{\dagger}$ | 82.1%$^{\ddagger}$ | 80.6%$^{\ddagger}$ | 4.35% | 4.25% | 10.30%$^{\ddagger}$ | 10.90%$^{\ddagger}$ |
| HILMA | 61.05%$^{\ddagger}$ | 65.15%$^{\ddagger}$ | 100%$^{\ddagger}$ | 100%$^{\ddagger}$ | 1.95%$^{\dagger}$ | 4.70% | 4.00% | 4.00% |

$^{\dagger}$: conservative

$^{\ddagger}$: inflated

Table S1: Type I error results for count exposure. We consider four scenarios and two correlation settings ($\rho = 0, 0.5$) under the null.

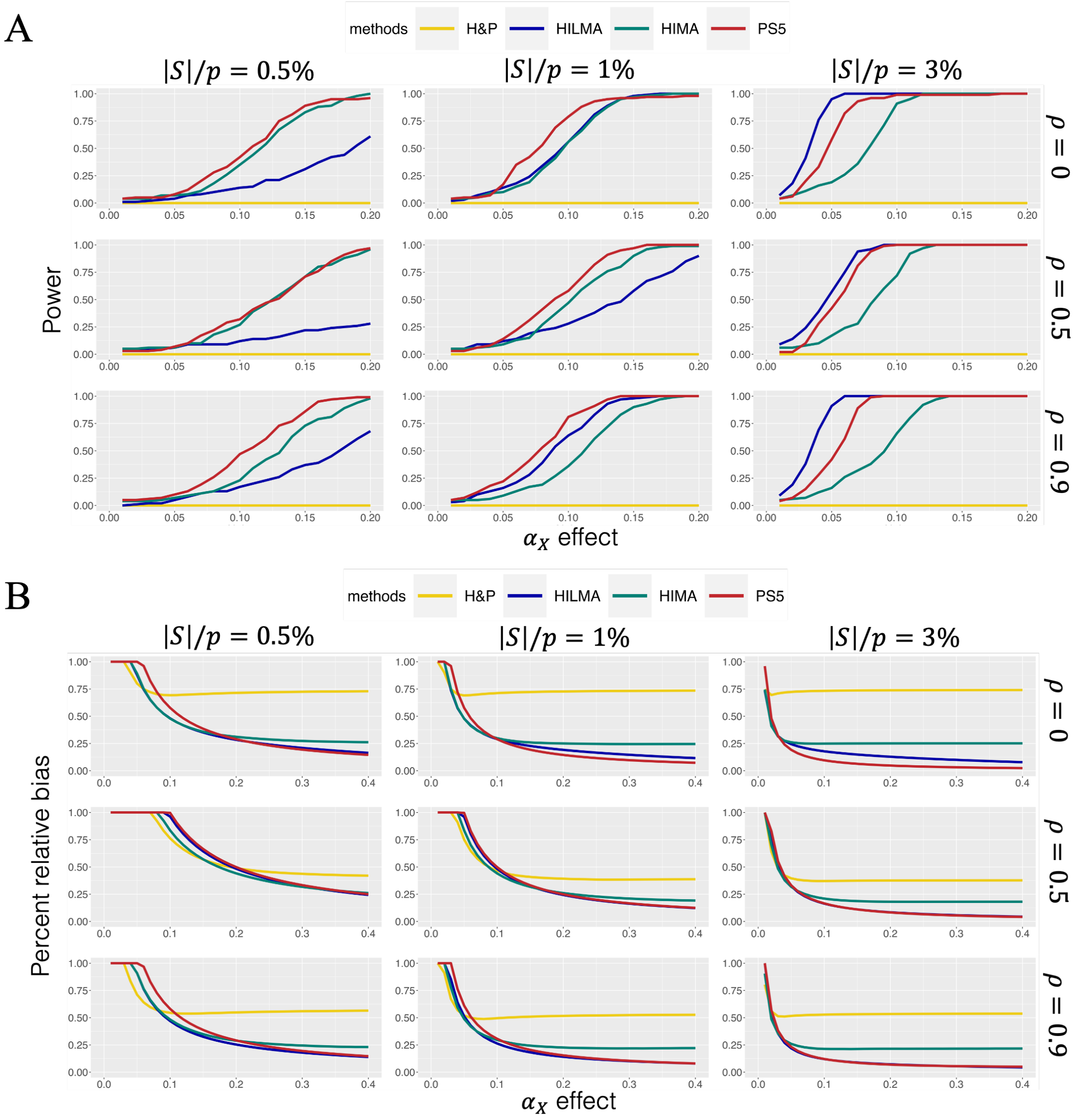


Figure S4: Simulation results for count exposure. (A) Power for detecting global indirect effect (B) Percent relative bias for estimating global indirect effect

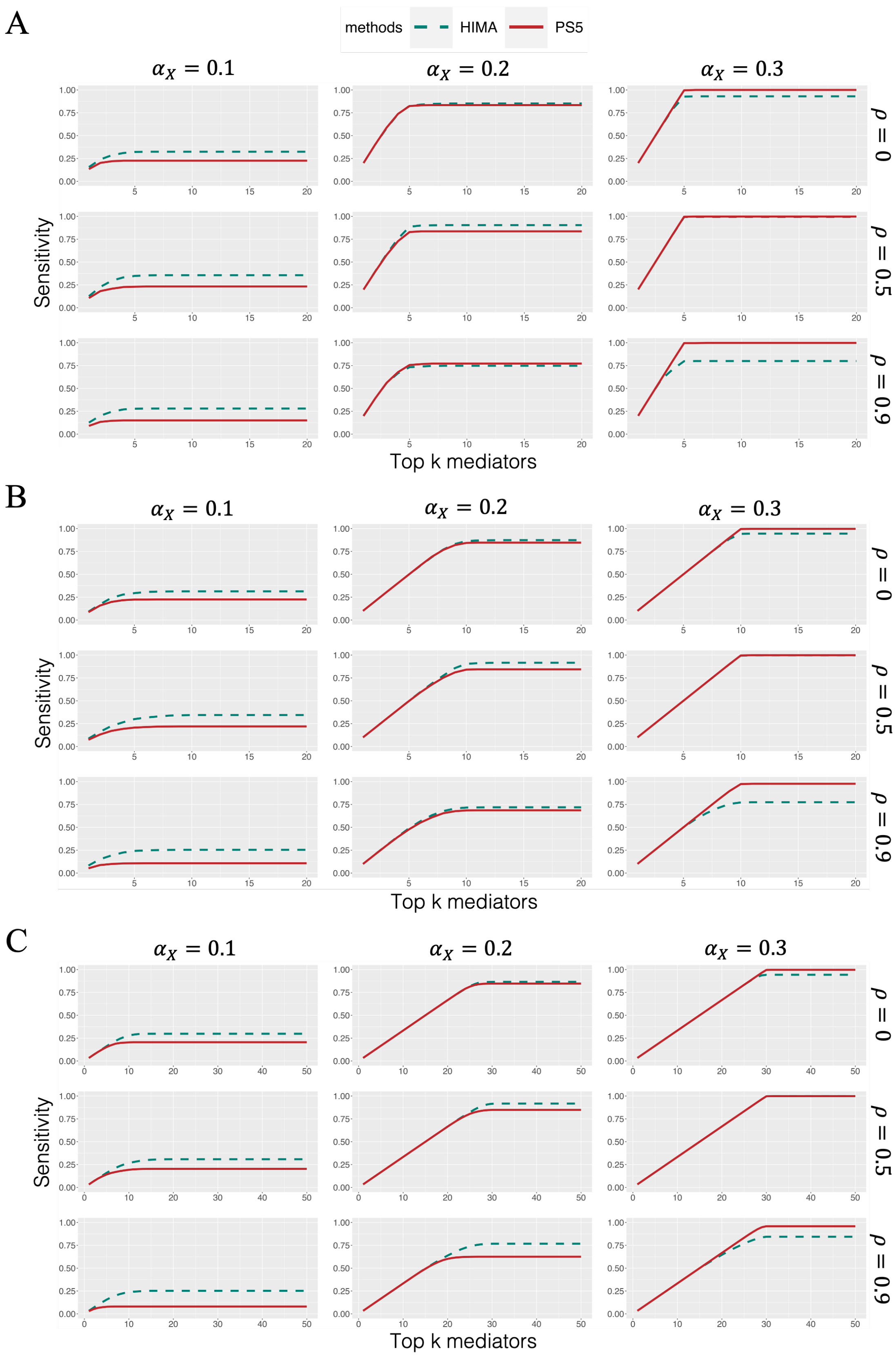


Figure S5: Accuracy of mediator prioritization for count exposure by sensitivity. (A) $|\mathcal{S}|/p = 0.5\%$ (B) $|\mathcal{S}|/p = 1\%$ (C) $|\mathcal{S}|/p = 3\%$
